# Suppression of Groups Intermingling as Appealing Option For Flattening and Delaying the Epidemiologic Curve While Allowing Economic and Social Life at Bearable Level During COVID-19 Pandemic

**DOI:** 10.1101/2020.05.06.20093310

**Authors:** Ioan Bâldea

## Abstract

In this work, we simulate the COVID-19 pandemic dynamics in a population modeled as a network of groups wherein infection can propagate both via intra-group and via inter-group interactions. Our results emphasize the importance of diminishing the inter-group infections in the effort of substantial flattening and delaying of the epi(demiologic) curve with concomitant mitigation of disastrous economy and social consequences. To exemplify with a limiting case, splitting a population into m (say, 5 or 10) noninteracting groups while keeping intra-group interaction unchanged yields a stretched epidemiologic curve having the maximum number of daily infections reduced and postponed in time by the same factor m (5 or 10). More generally, our study suggests a practical approach to fight against SARS-CoV-2 virus spread based on population splitting into groups and minimizing intermingling between them. This strategy can be pursued by large-scale infrastructure reorganization of activity at different levels in big logistic units (e.g., large productive networks, factories, enterprises, warehouses, schools, (seasonal) harvest work). Importantly, unlike total lockdwon strategy, the proposed approach prevents economic ruin and keeps social life at a more bearable level than distancing everyone from anyone.

## 1 Introduction

Neither natural immunization nor vaccination or pharmacologic intervention can currently help in fighting the COVID-19 global pandemic.^1^ Most frequently, to prevent disastrous sanitary consequences, governments across the world responded by imposing, e.g., social (=physical) distancing, wearing face masks, lockdown regulations, and rigid sanitary moves aiming at reducing the infection rate (“flattening the epidemiologic curve”). Still, politics cannot push strict restrictions indefinitely, and “how much is too much?” is a question of time which unavoidably arises sooner or later. Fighting COVID-19 should not ruin economy. ^2^ This is certainly what a draconian lockdown across the world over months would do. Parenthetically, catastrophic economic consequences inherently make healthcare system itself also collapsing.

**Figure 1:**
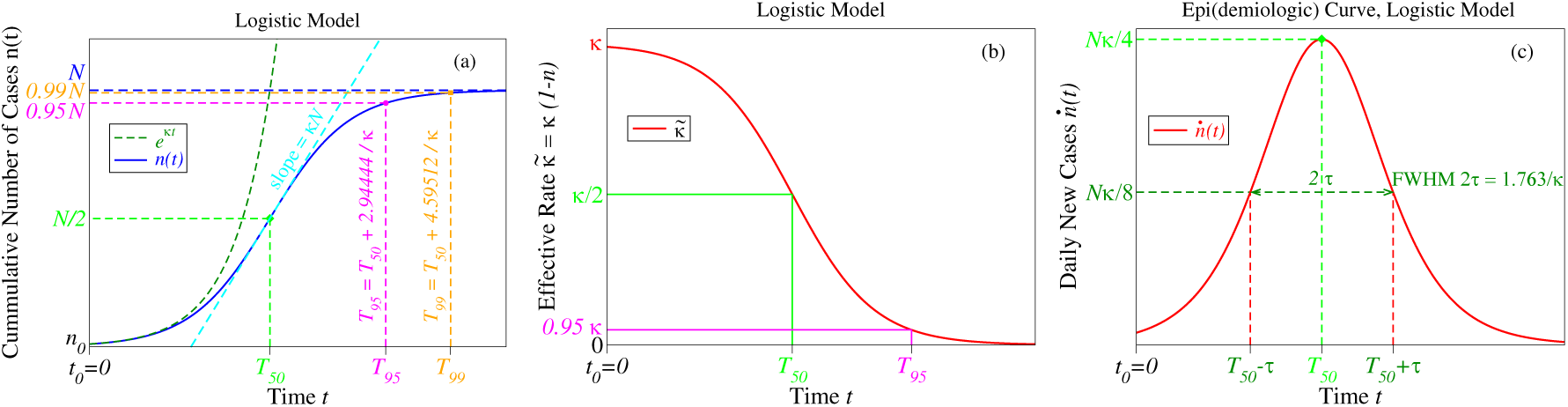
Logistic growth is characterized by a cumulative number of infected cases *n*(*t*) following an S-shaped (sigmoid) curve (panel a) exhibiting an acceleration stage (close to a J-shaped exponential growth), which switches to a deceleration stage beyond the half-time *T*_50_ and attains *p*(= 95, 99, see equation (12)) percent of the plateau value *N*. Saturation occurs because the effective infection rate 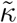 is time dependent and gradually decreases to zero (panel b). The epi(demiologic) curve *ṅ*(*t*) = *dn*/*dt* (daily number of new cases) exhibits a peak located at *t* = *T*_50_ whose shape is controlled by the infection rate *κ* (panel c). Time on the *x*-axis is expressed in units of the characteristic time *T_c_* = *κ*^−1^.

In this vein, mathematical modeling may make a notable contribution in providing politics with reasonable suggestions to slowing down epidemic propagation and reducing medical burden while mitigating economy and social crisis. A series of mathematical COVID-19 simulations have recently appeared.^3–10^ Most of them are based on deterministic continuous-time epidemiologic models, which consider age-independent epidemiologic classes of, e.g., susceptible (S), exposed (E), infected (I), and recovered (R) individuals,^11–16^ whose numbers S(t), E(t), I(t), R(t) evolve in time (t) according to a system of (deterministic) ordinary differential equations.

Open access sources already available^17–19^ enable one to easily perform various numerical simulations by means of such models. Unfortunately the various SIR-inspired flavors need (too many) input parameters difficult to validate,^20^ and this would rather mask than enlighten the main idea which the present work aims at conveying. Therefore, to better emphasize this idea, instead of a SIR-based approach (which would pose no special problem), in this paper we prefer to adopt the simpler logistic growth framework. The logistic model is particularly appealing in view of its simplicity and versatility demonstrated in approaching a broad variety of real systems with very different nature,^21–28^ including population dynamics of epidemic states.^29–32^ Prior to this study, results based on the logistic model were presented for COVID-19 time evolution in China and USA.^33^

Fighting against the spread of SARS-CoV-2 virus while allowing economic and social activity to continue to a reasonable extent represents a major challenge for the present era. Extended lockdown does not represent an acceptable response to this challenge. From this perspective, we believe that the results reported below obtained by extending the conventional logistic model may provide useful suggestions on how to sidestep the ongoing difficulty of living under pandemic conditions.

While the implementation of the presently proposed strategy via population splitting into smaller groups and reducing intermingling certainly requires considerable effort and fantasy in infrastructure reorganization, it offers the perspective of flattening and delaying the epidemiologic curve by obviating wrecking of economy and maintaining social life to a level more bearable than total lockdown. It can also be pursued as a complementary approach to the massive COVID-19 testing proposed to fostering economy recovery.^34^

## 2 Methods

The results reported below were obtained by means of the logistic model^21–28^ extended (see equation (16)) to allow treatment of infection propagation in a network consisting of groups in interaction. To fix the ideas and to make the paper self-contained, in Section 3.1 we will first review the main aspects related to the logistic model applied to an isolated group using a terminology adapted to the specific subject under consideration. The extension of the logistic model to groups in interaction will be presented in Section 3.2.

## 3 Results and Discussion

### 3.1 Logistic Growth in an Isolated Group

Uninhibited infected population n growths in time t according to the Malthus law^35^

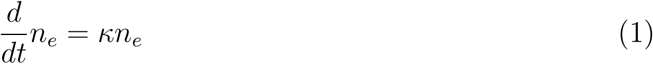

The intrinsic population-independent rate κ entering equation (1) is expressed in terms of the probability *β* of infection per encounter with an infected individual multiplied by the number 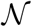 of encounters per unit time (day)

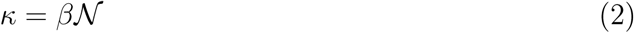

This yields an unlimited exponential time growth (*n*_0_ = *n* (*t* = 0))

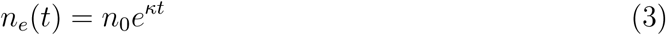

depicted by the dark green J-shaped curve of Figure 1a.

In a real situation, the exponential growth will gradually slow down and eventually level off. Infections become more and more unlikely because, in a given environment, the increase in the number of infected diminishes the number of individuals that can be infected. Rephrasing, the effective growth rate decreases with increasing population density: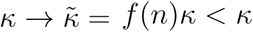. By assuming a linear decrease of *f* with *n*, one arrives at the logistic model^21–26^

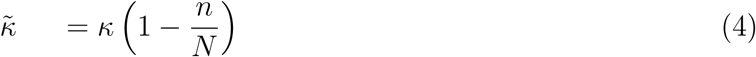

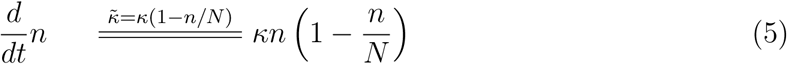

Plotted as a function of time (Figure 1a), equation (5) yields an exponential J-shaped curve only at early times which switches to an S-shaped (sigmoid) curve as the population increases and saturates to the maximum (plateau) value N, which defines the so-called carrying capacity of a given environment. The parenthesis entering the right hand side of equation (5), which acts as Darwin’s “struggle for existence” and suppresses the exponential growth, is similar to the Pauli blocking factor extensively discussed in electron transport theory.^36–40^

The continuous time representation underlying the differential equation (5) allows to express the cumulative number of cases *n*(*t*) in closed analytical form

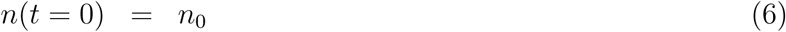

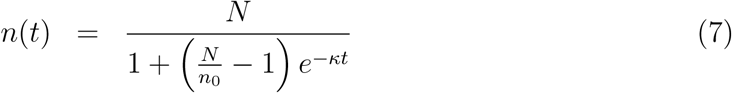

By using the half-time *T*_50_

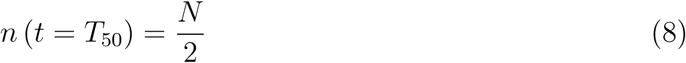

which defines the crossover point where the population attains *p* = 50 percent of its maximum (*N*), the above results can be recast as follows

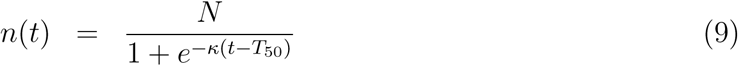

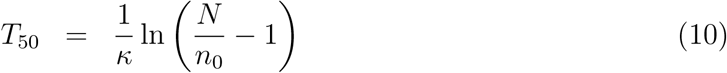

At *t* = *T*_50_, there is a substantial infection slowing with respect to the exponential growth. There, the instantaneous infection rate 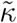 is reduced by 50% as compared to that of the uninhibited growth *κ* (equation (4) and Figure 1b).

Saturation occurs within a few characteristic times *T_c_* ≡ *κ*^−1^ beyond the half-time *T*_50_ (Figure 1a). The moment (“day”) *T_p_* when the number *N_p_* of infected amounts to *p* percent (e.g., *p* = 95 or 99, cf. Figure 1) of the maximum value *N*

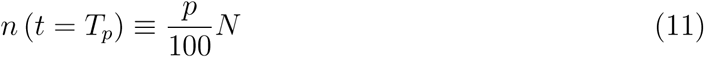

can easily be deduced from equation (9)

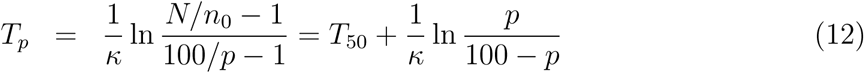

The quantity *κ* is important because it quantifies the daily new infected cases expressed by the time derivative *dn/dt* (equation (13) and the red curve in Figure 1c)

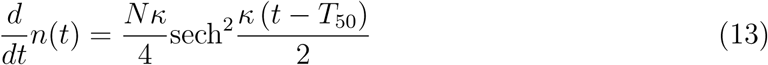

The height *H* and full width at half maximum FWHM of this so-called epidemiologic curve *dn*/*dt* = *f* (*t*) are expressed by

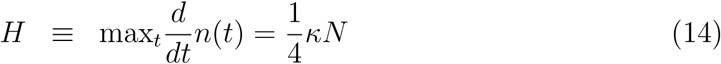

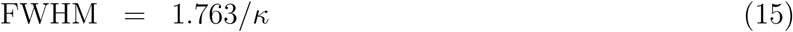

Large values of *κ*, amounting to a sharp and high peak in Figure 1c, may cause healthcare systems collapses.

Diminishing the value of *κ* is of paramount practical importance for a twofold reason:

i. It renders the peak broader and smaller (Figure 1c). Keeping the daily number of infected cases at a manageable level (“flattening of the curve”) is essential for not overwhelming the healthcare system beyond its capacity to treat the sick.
ii. Small values of κ yield large values of *T*_50_ (cf. equation (10)). This means postponement of infection explosion and hence gaining time for a better sanitary and logistic preparation to tackle an upcoming problem: preventing shortage of intensive care unit beds and gaining time for securing and/or producing critical emergency equipment (e.g., masks, ventilators, artificial lungs, personal protective equipment, extracorporeal membrane oxygenation machines, ventilators or other devices) needed to reduce mortality rate.

### 3.2 Modeling Infection in Interacting Groups

The above considerations referred to a closed population in which members are neither added nor lost from the group. Neither “imported” nor “exported” infections were included. Let us now focus on a network consisting of groups of individuals {*n_j_*} = {*n*_1;_ *n*_2_,…, *n_m_*} wherein infections can proliferate both by infections within the same group (intra-group infection rates *κ_j_* ≡ *κ_jj_*) and because individuals of one group j can infect or can be infected by individuals of other groups *p* ≠ *j* (inter-group/intermingling infection rates *κ_pj_* and *κ_jp_*, respectively). Figure 2 schematically depicts the case of two groups.

**Figure 2:**
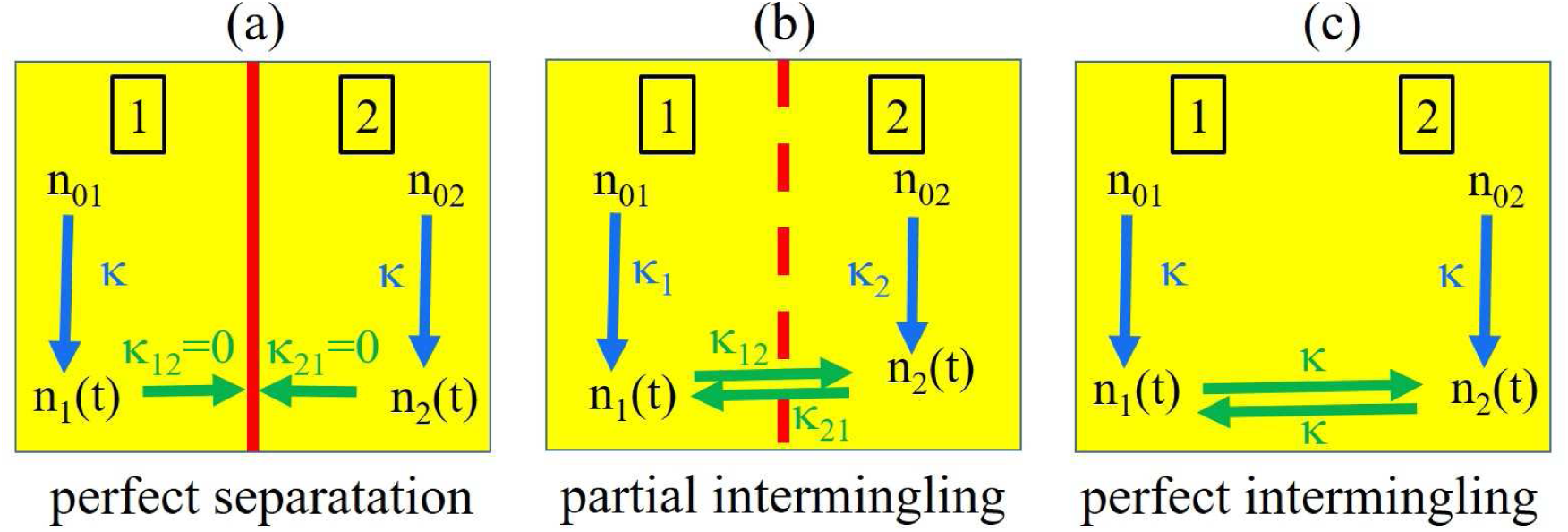
Schematic representation of a network consisting of two groups 1 and 2 wherein epidemic can spread (panel b) both through intra-group infections (infection rates *κ*_1_ and *κ*_2_) and through inter-group infections (infection rates *κ*_12_ and *κ*_21_). Limiting cases wherein the network is split into two groups that are completely separated among themselves *(κ*_12_ = *κ*_21_ = 0, panel a) or perfectly intermingled (*κ*_12_ = *κ*_12_ = *κ*_21_ = *κ*, panel c).

Generalizing the idea underlying equation (5), we will consider below the following extended logistic model

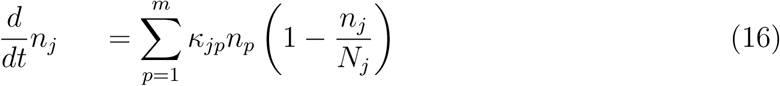

Although solving equation (16) in general poses no special numerical problem (some examples are presented in Figure 4 and Figure 5 of Section 3.4), to better emphasize the strategy we aim at conveying, let us first focus on the case of identical groups

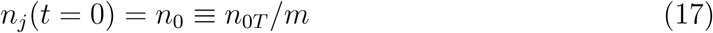

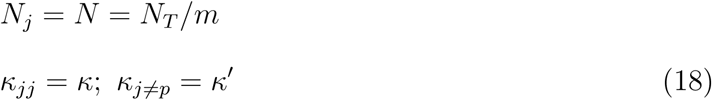

Equation (18) yields *j*-independent populations

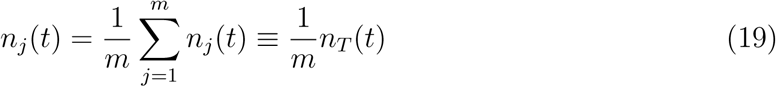

*n_T_* (*t*) being the total time dependent population.

Upon term-by-term addition (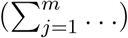) of equation (16) we immediately get

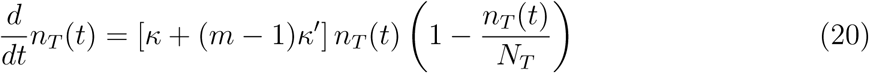

and hence

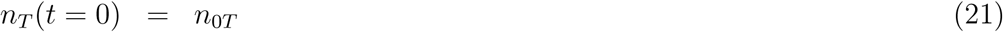

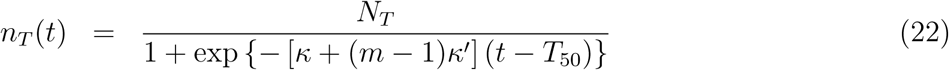

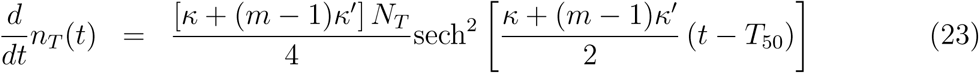

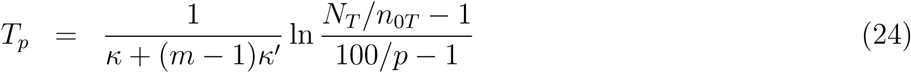

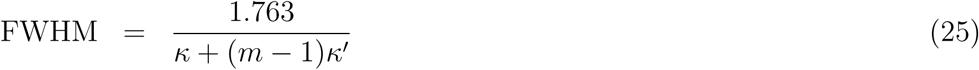

By comparing the above formulas with equations (6)-(10) valid for a single group one can conclude that the quantity

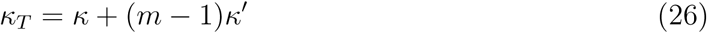

plays the role of a total infection rate. Importantly, both the half-time *T*_50_ and the maximum number of daily infections *H* deduced from equations (23) and (24)

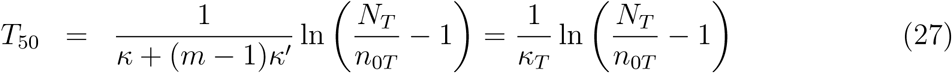

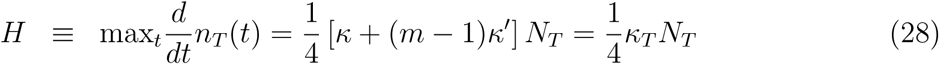

are controlled by κ*_T_*.

### 3.3 Analysis of Two Limiting Cases of Practical Importance

The results of Section 3.2 allow us to compare how infection propagates in a network (“larger group”) of individuals split into several (*m*) smaller (sub)groups (chosen identical for simplicity) which do not interact with each other against the case of a fictitious splitting, wherein (from the point of view of infection) interactions between members of a given group and between members belonging to different groups are identical (perfect intermingling). Noteworthy, whether the groups are completely separated of each other (label *s*) or perfectly intermingled (label *i*), the total initial population *n*_0_*_T_* = *mn*_0_ is taken to be the same in both cases (cf. equation (17))

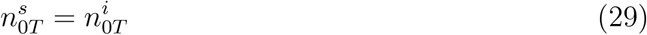

The former case, corresponding to separated groups (label *i*), is characterized by a vanishing inter-group infection rate (*κ*′ ≡ 0). Applied to this case, equations (26), (27) and (28) yield

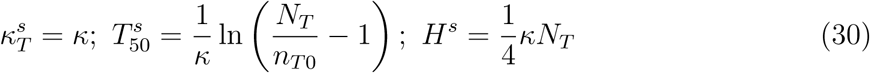

At the opposite extreme of perfectly intermingled groups, inter-group interactions are as strong as intra-group interactions (*κ*′ = *κ*). Based on equations (26), (27) and (28) we then get

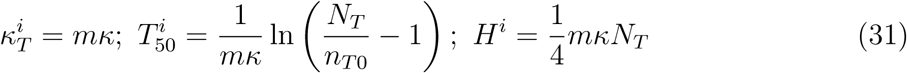

The above results show that the epidemiologic curve can be substantially flattened if a larger group is split into several smaller groups separated from each other. By starting from the same number of infections (equation (29)), splitting into groups separated from one another yields a reduction of the infection rate and of the maximum daily cases by a factor *m* and an increase of full width at half maximum by the same factor

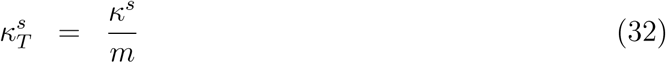

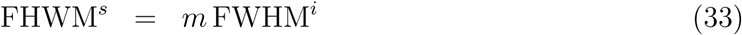

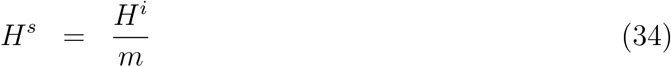

Equally pleasantly, splitting leads in addition to a time postponement of the infection peak by the same factor *m*

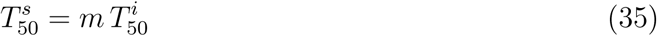

The results presented in Figure 3 depict these findings for the particular case of a larger group split into two smaller groups (*m* = 2). They also schematically visualize how and why group splitting can relieve the healthcare system. To avoid misunderstandings, one should note that the value *m* = 2 in Figure 3 was chosen just for more clarity. Splitting into more than two noninteracting groups (i.e., making *m* as large as possible) is highly desirable.

**Figure 3:**
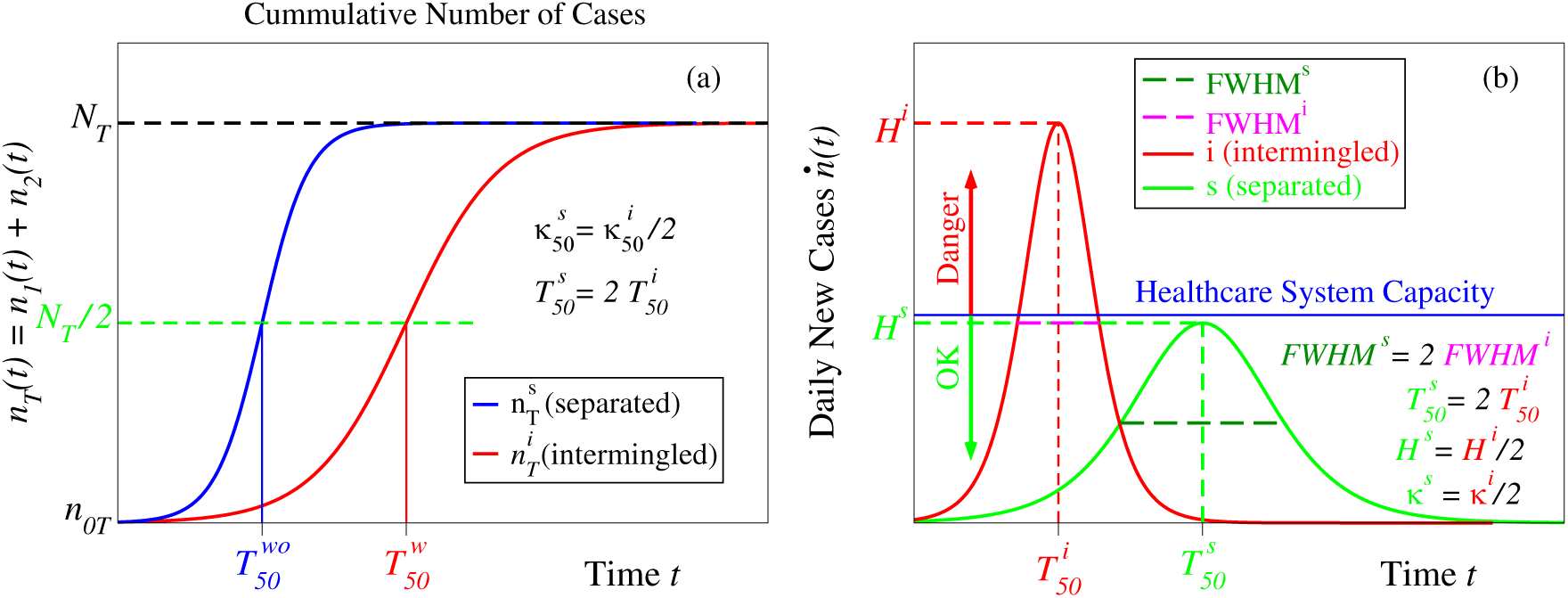
Results demonstrating how preventing intermingling between several groups flattens the epidemic curve with the concomitant time delay of the maximum of daily infections. (a) Cumulative number of cases *n*_1,2_(*t*) and (b) epidemiologic curve *ṅ*_1,2_(*t*) ≡ *dn*_1,2_*(t)* for a group consisting of two subgroups that are either completely separated of each other (green lines) or perfectly intermingled (red curves). These results also schematically depict how flattening and delaying the epidemiologic curve by a factor *m* = 2 by splitting into *m* = 2 groups can prevent overloading the healthcare system capacity. Time on the *x*-axis is expressed in units of the characteristic time *T_c_* = κ^−1^.

Before ending this part, we want to emphasize that, however important, flattening and delaying the epidemiologic curve by a factor m achieved by group splitting (cf. Figure 3 and equations (32), (33), and (34)) is not the whole issue. Extremely importantly, the presently proposed group splitting approach does not assume any intra-group (like social distancing and wearing masks) restrictions: economy and social life within individual groups separated of each other can continue.

### 3.4 Additional Results

As anticipated in Section 3.2, the logistic model extended as expressed by equation (16) can be used to quantify the impact of mutual infections in networks of interacting groups more general than the particular situations examined in Section 3.3. To briefly illustrate this fact, two examples are presented in Figures 4 and 5.

**Figure 4:**
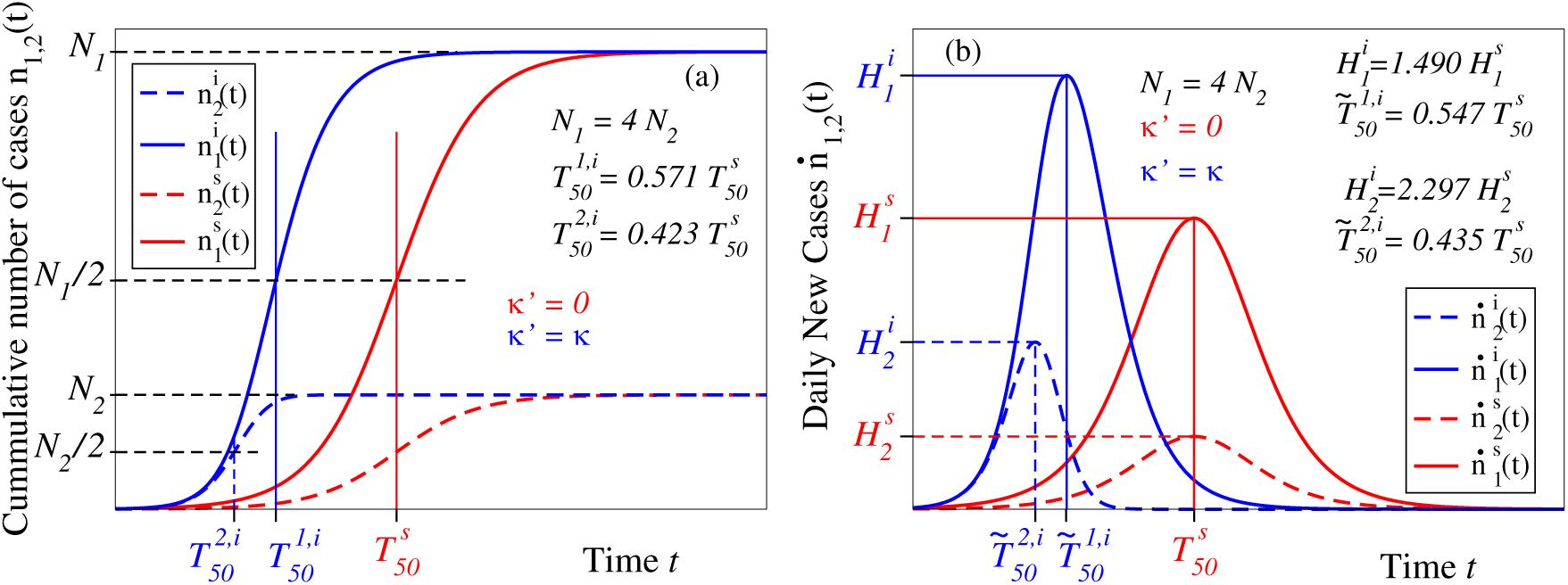
(a) Cumulative number of cases *n*_1,2_ (*t*) and (b) epidemiologic curves *ṅ*_1,2_(*t*) = *dn*_1,2_(*t*)/*dt* for two completely separated and perfectly intermingled (red and blue curves, respectively) groups whose populations differ by a factor of four. Notice that, in spite of the equal infection rates, infection of the smaller group 2 (dashed lines) is stronger affected by the inter-group interaction than in the larger group 1 (solid lines). Time on the x-axis is expressed in units of the characteristic time *T_c_* = *k*^−1^.

**Figure 5:**
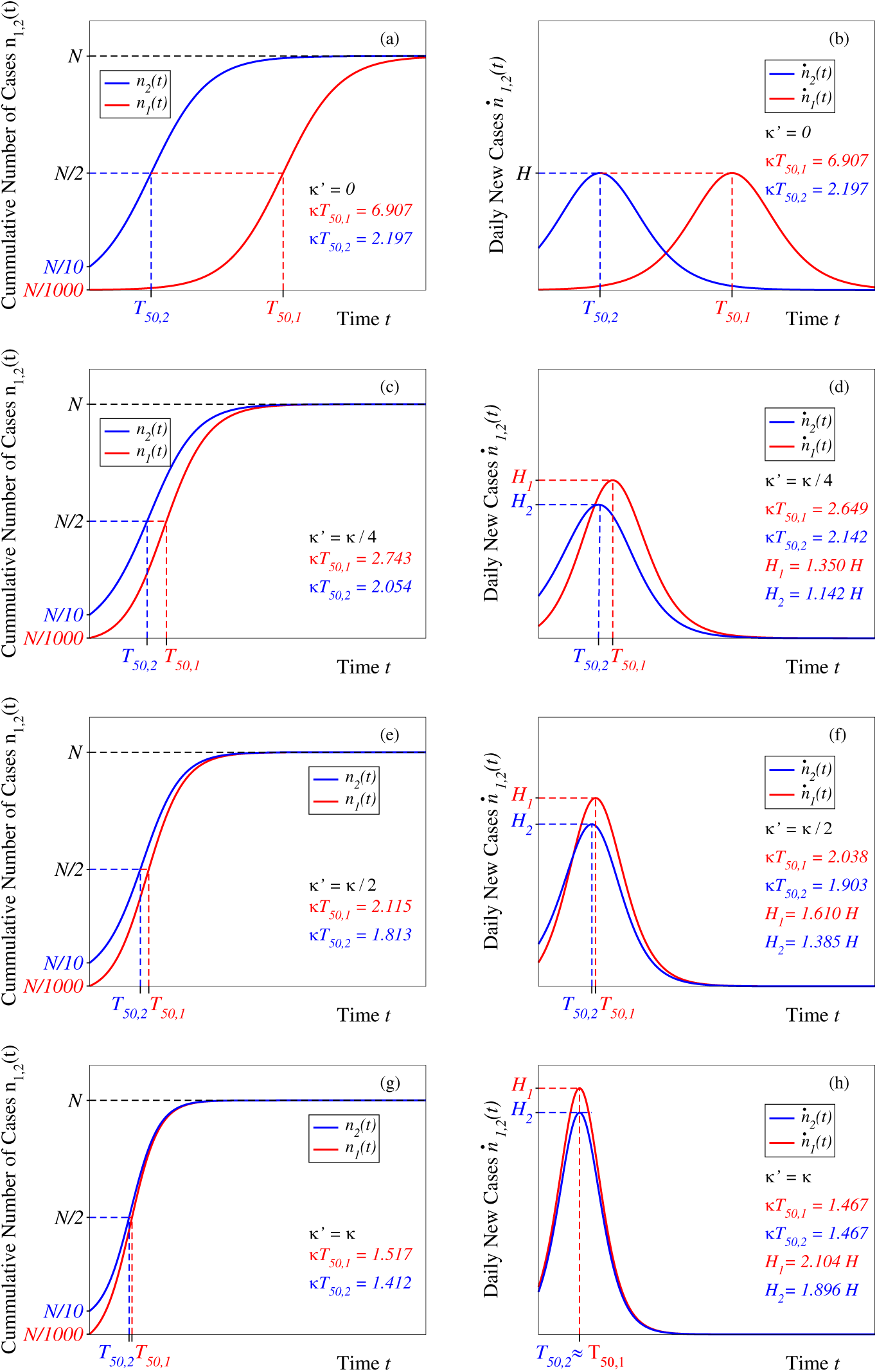
Results showing the impact of the inter-group interaction *K* on the cumulative number of cases *n*_1,2_(*t*) and epidemiologic curves *ṅ*_1,2_(*t*) = *dn*_1,2_(*t*)/*dt* in case of two groups merely differing by the initial number of infected cases: *n*_01_/*N* = 0.001 and *n*_02_/*N* = 0.1. They show how infections in the initially less infected group 1 are rapidly triggered by infections in the initially more infected group 2. Time on the *x*-axis is expressed in units of the characteristic time *T_c_* = *κ*^−1^.

Figure 4 depicts the case two groups whose populations differ by a factor of four. As visible there, in spite of the equal infection rates, infection of the smaller group 2 (dashed lines) is stronger enhanced by the inter-group interaction than in the larger group 1 (solid lines).

Figure 5 presents the case two groups merely differing from each other by the different numbers of initially infected individuals *(n*_01_*/N* = 0.001 versus *n*_02_/*N* = 0.1). Comparison of the various panels of Figure 5 reveals that inter-group infection yields an infection rapidly “exported” from the initially more infected group to that which was initially less infected. Intermingling (Figure 5g) quickly wipes out any difference between a initially weaker (or non)infected group and an initially strongly infected group.

## 4 Conclusion

In closing, the results presented above indicate that splitting of a large group into smaller groups and reducing intermingling appears to be an appealing strategy for substantially reducing the spread of the SARS-CoV-2 virus while still allowing social life to a more bearable level than distancing everyone from anyone and economy go on, albeit slower.

One can expect that (A) it is easier to impose and maintain longer-term regulations on splitting a given population into (say, 5 – 10) groups weakly interacting among themselves than (B) enforcing severe containment measures to all individuals in order to diminish the probability *β* of infection per encounter and the number 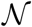 of daily encounters yielding a reduction by the same factor (5–10) of the infection rate κ (cf. equation (2)). The big difference is that with option (A) social life and economy within individual groups go on without intra-group restrictions, while option (B) means attaining the same epidemiologic curve with both social life and economy paralyzed.

Devising and implementing an adequate restructuring of large logistic units (large productive networks, factories, enterprises, warehouses, etc) allowing, if/when necessary, society to rapidly switch back and forth between separated groups and intermingled groups can certainly be challenging but may be a long-term strategic goal worth to be pursued when faced with these COVID-19 pandemic times or other similar difficulties that cannot be ruled out in the future.

## Data Availability

The manuscript includes all information necessary to generate and reproduce the data reported in the present work.

